# Inter-individual differences in the blood pressure lowering effects of dietary nitrate: A randomised double-blind placebo-controlled replicate crossover trial

**DOI:** 10.1101/2024.12.02.24318286

**Authors:** Eleanor Hayes, Shatha Alhulaefi, Mario Siervo, Eleanor Whyte, Rachel Kimble, Jamie Matu, Alex Griffiths, Marc Sim, Mia Burleigh, Chris Easton, Lorenzo Lolli, Greg Atkinson, John C Mathers, Oliver M Shannon

## Abstract

**Purpose:** Dietary nitrate supplementation increases nitric oxide (NO) bioavailability and reduces blood pressure (BP). Inter-individual differences in these responses are suspected but have not been investigated using robust designs, e.g., replicate crossover, and appropriate statistical models. We examined the within-individual consistency of the effects of dietary nitrate supplementation on NO biomarkers and BP, and quantified inter-individual response differences.

**Methods:** Fifteen healthy males visited the laboratory four times. On two visits, participants consumed 140ml nitrate-rich beetroot juice (∼14.0mmol nitrate) and, on the other two visits, they consumed 140ml nitrate-depleted beetroot juice (∼0.03mmol nitrate). Plasma nitrate and nitrite concentrations were measured 2.5 hours post-supplementation. BP was measured pre– and 2.5 hours post-supplementation. Between-replicate correlations were quantified for the placebo-adjusted post-supplementation plasma nitrate and nitrite concentrations and pre-to-post changes in BP. Within-participant linear mixed models (LLM) and a meta-analytic approach estimated participant-by-condition treatment response variability.

**Results:** Nitrate-rich beetroot juice supplementation elevated plasma nitrate and nitrite concentrations and reduced systolic (mean:-7mmHg, 95%CI: –3 to –11mmHg) and diastolic (mean:-6mmHg, 95%CI: –2 to –9mmHg) BP *versus* placebo. The LLM participant-by-condition interaction response variability was ±7mmHg (95%CI: 3 to 9mmHg) for systolic BP and consistent with the treatment effect heterogeneity τ=± 7mmHg (95%CI: 5 to 12mmHg) derived from the meta-analytic approach. The between-replicate correlations were moderate-to-large for plasma nitrate, nitrite and systolic BP (r=0.55 to 0.91).

**Conclusions:** The effects of dietary nitrate supplementation on NO biomarkers and systolic BP varied significantly from participant to participant. The causes of this inter-individual variation deserve further investigation. Trial registration: https://clinicaltrials.gov/study/NCT05514821.

## INTRODUCTION

High blood pressure (BP) is a leading cause of morbidity and mortality worldwide [1], and the identification of effective strategies to lower BP remains a major research and public health priority [2, 3]. Consumption of a healthy diet can lower BP and is recommended as a first line treatment for low grade hypertension and as an adjunct to BP-lowering pharmacotherapy for more severe hypertension [4]. Various dietary approaches have been demonstrated to help lower BP, including consumption of dietary compounds/foods with potential anti-hypertensive properties such as dietary inorganic nitrate [5–7].

For most people, the major dietary sources of inorganic nitrate are vegetables such as lettuce, spinach and beetroot [8, 9], and this nitrate serves as a precursor for nitric oxide (NO) – a gaseous signalling molecule with vasodilatory properties [10, 11]. Consumption of nitrate-rich vegetables or vegetable juices (e.g., [6, 12–14]), and nitrate salts (e.g., [5, 15, 16]), has been shown to elevate NO biomarkers (e.g., plasma nitrate and nitrite concentrations) and lower BP both acutely (within hours of supplementation) and chronically (over several weeks/months) [7]. However, individual differences in the response to nitrate have been suggested, with some researchers hypothesising the existence of individuals deemed ‘*responders*’ and ‘*non-responders*’ to nitrate supplementation [17–21]. If correct, this notion could have important implications for the development of personalised recommendations around nitrate intake. For example, if it is possible to identify the individuals most likely to benefit from nitrate supplementation, then these individuals could be targeted for nitrate-based interventions in future trials and public health initiatives.

Identification of meaningful inter-individual differences in response to nutritional interventions can be challenging. Notably, with conventional parallel-arm or crossover trials, it is not possible to determine whether any apparent difference between participants in the changes from pre-to post-intervention are due to genuine treatment response heterogeneity or are a consequence of random within-subject variability and/or measurement error [22]. Without repeated administration of trials, a typical crossover design does not allow formal estimation of variance attributable to the participant-by-treatment interaction [23, 24]. Only with knowledge about this interaction can treatment response heterogeneity be quantified properly. As a form of n-*of*-1 trial, a replicate crossover trial constitutes a pragmatic research design for quantifying treatment response heterogeneity [22, 23, 25]. This research design involves repeated administration of the intervention and control/placebo arms of a trial protocol in randomised order on at least two occasions. This allows quantification of treatment response heterogeneity using, for example, a within-participant covariate-adjusted linear mixed model for estimation of any participant-by-treatment interaction effects [22, 23, 25].

We aimed to quantify the magnitude of inter-individual variability in the effects of dietary nitrate supplementation on the outcomes of NO biomarkers and BP using a replicate crossover design. We also aimed to examine the consistency of these responses on repeated occasions. We hypothesized that there is ‘*true*’ inter-individual variability (i.e., exceeding random within-subject variability) in the effects of dietary nitrate supplementation on NO biomarkers and BP, and that these responses would be consistent on two occasions.

## METHODS

The protocol for this study was registered prospectively on ClinicalTrials.gov (NCT05514821). The study was approved by the Faculty of Medical Sciences Research Ethics Committee at Newcastle University (2345/23609).

### Participants

Healthy male participants were recruited from the general population via posters, university email lists, and social media to take part in this study (see Supplementary Figure 1 for CONSORT flow chart). Participants were required to be non-smokers who were not currently taking any medication or using any dietary supplements, had no history of cardiovascular, metabolic, or gastrointestinal diseases, and were not currently using antibacterial mouthwash.

### Design

Participants attended the laboratory on 5 separate occasions (data collection period: October 2022 to September 2023). On the first visit, participants provided written informed consent and underwent screening to determine eligibility to participate. A pre-screening questionnaire was completed, and body mass and stature were measured. Subsequently, participants completed four experimental visits (two nitrate and two placebo) in a randomised order. A randomised sequence schedule (see Supplementary Text 1) was created using http://www.randomization.com) for our replicate crossover experimental research design [25]. Experimental visits were separated by ∼7 days (minimum 3 days, maximum 14 days). Participants were asked to record their diet in the 24 hours prior to the first visit and to replicate this as closely as possible prior to each subsequent visit. Participants were asked to abstain from intensive exercise and alcohol in the 24 hours prior to each visit and were instructed to avoid consumption of any food or drink except for plain water on the morning on the experimental visits.

### Experimental visits

Participants arrived at the laboratory between 8 and 9 am and rested, seated in quiet room for 10 minutes. Subsequently, systolic and diastolic BP of the brachial artery was measured using an automated sphygmomanometer following best-practice guidelines [26]. Four measurements were taken, with the mean of the final three measurements used for subsequent analyses. Participants then received a standardised breakfast including 140 ml concentrated nitrate-rich (∼14.0 mmol nitrate) or nitrate-depleted (∼0.03 mmol nitrate) beetroot juice and a bowl of porridge (60g oats, 200 ml whole milk). We used commercially available beetroot juice supplements (Beet It Sport, James White Drinks Ltd., Ipswich, UK). All supplements were from the same batch, with each batch undergoing homogenisation during manufacturing to maximise consistency in the nitrate concentrations. We analysed a single bottle each of the ‘active’ and placebo supplement using the same chemiluminescence approach outlined below to provide indicative nitrate concentrations. Supplements were administered double blind.

Participants then rested, seated in a quiet room for 2.5 hours during which time they were permitted to carry out non-stimulating activities (e.g., reading). Blood pressure measurements were then repeated, and a blood sample was collected via venepuncture of an antecubital vein into two, 4 ml lithium heparin containing tubes (Vacutainer, Becton Dickinson, Plymouth, UK). Samples were immediately centrifuged at 3000 rpm and 4°C for 10 minutes, plasma was extracted and frozen at –80℃. Participants were then free to leave the laboratory.

### Biochemical analyses

Measurements of plasma nitrate and nitrite concentrations were conducted using ozone-based chemiluminescence [27]. For the measurement of plasma nitrate concentration, vanadium reagent (24 mg of vanadium tri-chloride and 3 ml of 1M hydrochloric acid) and 100 μL of anti-foaming agent were placed into a glass purge vessel infused with nitrogen and heated to 95°C. This purge vessel was connected to a NO analyser (Sievers NOA 280i, Analytix, UK). A standard curve was produced by injecting 25 μL of nitrate solutions (100 μM, 50 μM, 25 μM, 214 12.5 μM, and 6.25 μM) and a control sample containing deionised water. The area under the curve (AUC) for the control sample was subtracted from those for the nitrate solutions to account for nitrate in the water used for dilutions. Plasma samples were thawed in a water bath at 37°C for 3 min and de-proteinised using zinc sulphate/sodium hydroxide solution (200 μL of plasma, 400 μL of zinc sulphate in deionised water at 10% w/v and 400 μL of 0.5M sodium hydroxide). The samples were then vortexed for 30 s before being spun at 4000 rpm for 5 min. Subsequently, 15-25 μL of the sample was injected into the purge vessel in duplicate. The concentration of NO produced was then measured by the NO analyser. The AUC was calculated using Origin software (version 7) and normalised using the Y value from the calibration curve.

For the measurement of plasma nitrite concentrations, 2.5 ml glacial acetic acid, 0.5 ml of 18 Ω deionised water, 25 mg sodium iodide, and 100 μL of an anti-foaming agent were placed into the glass purge vessel and heated to 50°C. A standard curve was produced by injecting 100 μL of nitrite solutions (1000 nM, 500 nM, 250 nM, 125 nM, and 62.5 nM) and a control sample of deionised water. The AUC for the latter was subtracted from those for the nitrite solutions to account for nitrite in the water used for dilutions. Following this, plasma samples were thawed in a water bath and 100 μL of the sample was injected into the purge vessel in duplicate. The nitrite concentration was determined via the AUC, as described for nitrate analysis.

### Statistical analysis

We adopted a pragmatic approach to sample size considerations [28, 29]. Given the onerous nature of the four-condition replicate design and procedures, we recruited 15 participants, which is a sample size similar to previous replicate crossover trials in nutrition research [30, 31]. As detailed below, the between-replicate correlation coefficient is an indicator of individual stability of response. Using GPower (version 3.1), we estimated the minimum statistically significant between-replicate correlation coefficient to be 0.44 for a sample size of 15. The 90% confidence interval (CI) for this correlation coefficient is estimated to be 0.01 to 0.74. A one-tailed directional hypothesis (90% CI) is relevant here because the null hypothesis (H_o_) is that r = 0 OR <0, i.e., if the correlation is either zero or negative, this would lead to the same conclusion (non-rejection of H_o_) of no consistent responses being present [32].

The analysis protocol (which included data for all 15 participants) followed a four-step approach consistent with previous research [30, 31, 33] and more recent advances [34] relevant to the elaboration of continuous data from a replicate crossover trial designed experiment. Primary outcome measures were systolic and diastolic BP. Secondary outcome measures were plasma nitrate and plasma nitrite. First, we estimated Pearson’s product moment correlation coefficients between the first and second response replicates for each outcome to assess the within-person stability of the replicated placebo-adjusted supplementation effect [22] – a high correlation between the two repeated responses indicating a relatively stable individual response. For this correlation analysis of consistency of response, the first supplementation condition of each participant was paired to their respective first placebo condition in their individual sequence and the placebo-adjusted supplementation effect was computed for response 1 (supplementation 1 *minus* placebo 1 for BP changes). This process was replicated for the second condition pairs to calculate response 2 (supplementation 2 *minus* placebo 2).

Second, an overall “naive” estimate of the true individual difference standard deviation (SD) for the supplementation response was derived using the following approach [23]:

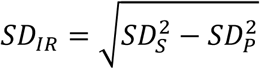

The SD*_IR_* value in the equation is the SD of the individual differences in the supplementation response (SD*_S_*) adjusted for the standard deviation in the placebo conditions (SD*_P_*), respectively [23]. A positive SD*_IR_* indicates that the variability in supplementation response is greater than any random within-subject variability. This calculation is considered a naive estimation of the SDir because it is not derived from a statistical model on the raw data and was originally formulated with parallel arm studies in mind. Specifically, our design is of a within-subjects nature rather than a parallel-arm study.

Third, and in view of the issues above with the naive estimate of SDir, we used the PROC MIXED procedure in SAS OnDemand for Academics (SAS Institute) to derive the SDir, in line with previous studies [23]. Distinct within-participant covariate-adjusted linear mixed models were used to quantify inter-individual differences in NO biomarkers and blood pressure responses specified as participant-by-condition interaction terms, with the variance-covariance matrix structure set to *variance components* [22, 25, 35]. Each model included the outcome measure specified as response variable, condition, period (condition sequence) and the period-by-condition interaction included as fixed effects, with participant and the participant-by-condition interaction modelled as random effects (Supplemental File 1). The variance for the participant by condition interaction indicates the degree of response heterogeneity and the P-value for this interaction represents whether the variance can be considered statistically significant from zero (no response heterogeneity). The adequacy of the modelled covariance parameter estimates was assessed via formal residual diagnostics procedures [36, 37]. Effects were reported as estimated marginal means alongside relevant measures of dispersion (SD) and uncertainty (95% CI). Using data from prognostic studies in cardiovascular medicine as a guide, we defined a Δ=2 mmHg reduction in systolic and diastolic BP as the minimal clinically important difference (MCID) when interpreting the meaningfulness of interindividual response differences [38, 39].

Fourth, according to Senn’s meta-analytical approach, a sample estimate of within-subjects variance was calculated and converted to a standard error using appropriate degrees of freedom to derive replicate-averaged treatment effects for each participant [34]. Using the *metagen()* function available in the *meta* package [40], we conducted a random-effects meta-analysis with Hartung-Knapp adjustment [41] to summarise individual-participant replicate-averaged treatment effects and respective sampling errors [34]. The restricted maximum-likelihood estimation method determined the tau-statistic (τ) value describing the between-participant replicate-averaged treatment effect response variability across the distribution of true treatment effects [42, 43]. As Senn [34] explained, there are typically few degrees of freedom per individual in an n-of-1 trial, so it is better to use a pooled variance in the meta-analysis, resulting in the same variance estimate being applied to each person in the case of a replicate crossover where the number of replicates is the same for each individual. The generalised Q-statistic method estimated the uncertainty surrounding the point τ-statistic value reported as 95%CI [44]. Weighted raw replicate-averaged treatment effect differences were presented as descriptive statistics together with the respective 95% prediction interval (95% PI) illustrating the range for the distribution of true mean differences expected for 95% of similar future studies [45, 46]. Meta-analyses were conducted in R (version 3.6.3, R Foundation for Statistical Computing); see Supplementary file 1 for the associated R Markdown file.

Additional exploratory analyses involved post-hoc correlations (Pearson’s correlation) estimated to examine relationships between plasma nitrite and nitrate with BP variables. Relationships were further examined using within-participant covariate-adjusted linear mixed effects models including the BP change score as dependent variable, plasma nitrite, condition, period, and the period-by-condition interaction as fixed effects plus a study participant random effect using the *xtmixed* command (StataMP v14.1; StataCorp LP, College Station, TX). Estimated marginal means described the expected BP changes at pre-specified plasma nitrite concentrations whose uncertainty was presented as 95%CI and derived using the *margins* commands [47].

## RESULTS

Fifteen healthy male participants with mean (SD) age of 27 (5) years and BMI of 24.0 (4.1) kg/m^2^ took part in this study.

### Plasma nitrate concentration

There was a strong positive correlation (r=0.91, 90%CI: 0.78 to 0.96) between the two sets of placebo-adjusted responses to nitrate supplementation for plasma nitrate concentration (Figure 1A). The within-trial SD for plasma nitrate concentration was substantially greater for the nitrate supplementation *versus* placebo conditions (Table 1). The model-based treatment-by-condition interaction response variability was ±45 uM (95%CI, 22 to 60 uM).

**Figure 1.**
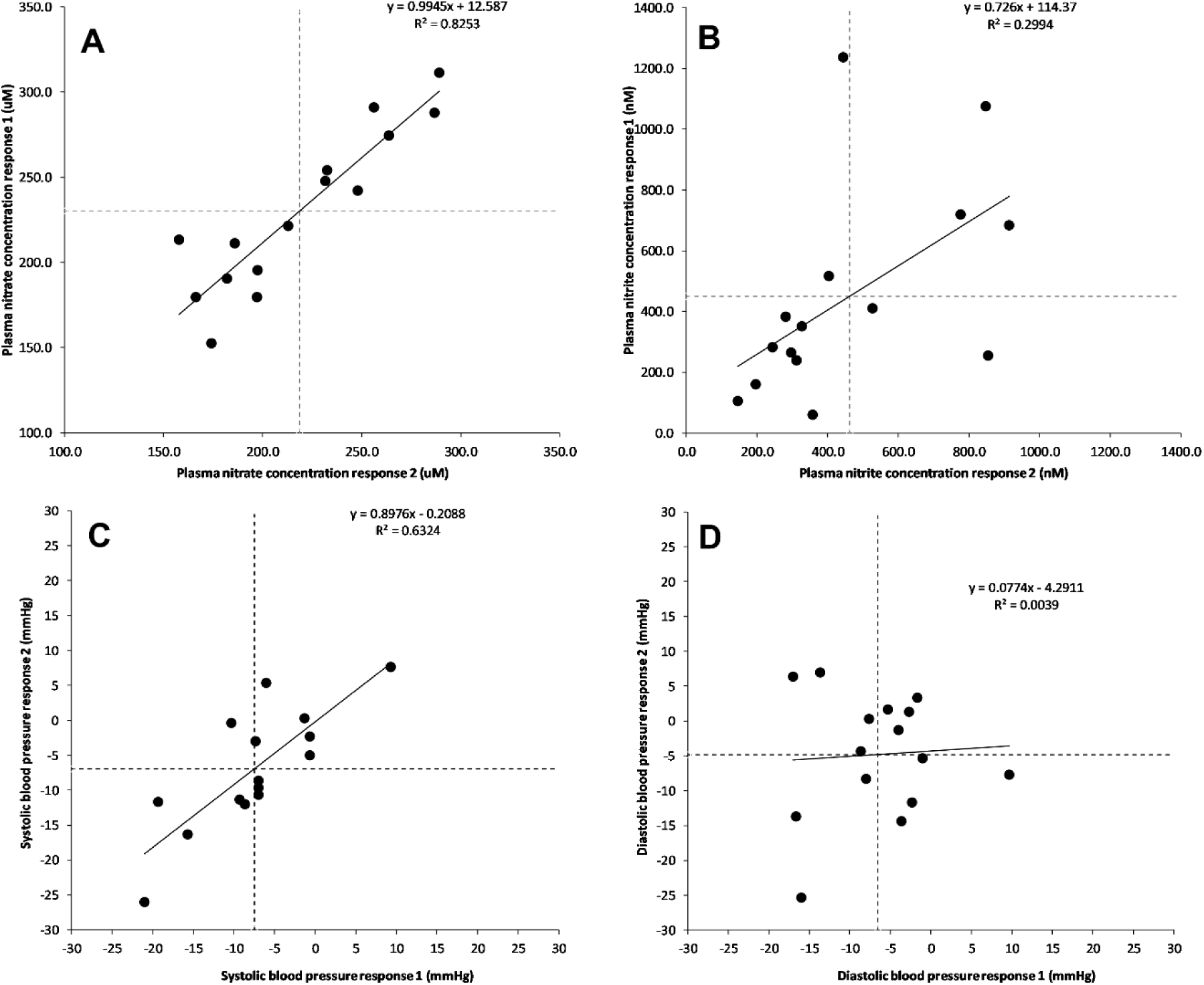
Individual panels present the relationship between the placebo-adjusted plasma nitrate concentrations (uM, panel A), plasma nitrite concentrations (nM, panel B), systolic BP (mmHg, panel C), and diastolic BP (mmHg, panel D) on the two occasions. “Response 1” reflects data for the first pair of conditions (nitrate 1 minus placebo 1) and “response 2” for the second pair of conditions (nitrate 2 minus placebo 2). The dashed vertical and horizontal lines reflect the mean responses.

**Table 1.** Estimated marginal means and SEs for primary outcome measures in the supplementation and placebo conditions with the true individual differences SD.

| Outcome | Mean (SE) |  | Main effect of condition | Estimate 1<sup>1</sup> | Estimate 2<sup>2</sup> | *P* value<br>(*int*) |
| --- | --- | --- | --- | --- | --- | --- |
|  | Supplementation | Placebo | Mean difference<br>(95% CI) | Individual<br>differences SD | Individual<br>differences SD<br>(95% CI) |  |
| Plasma nitrate concentration (uM) | 239 (12) | 15 (2) | 225<br>(199 to 250) | 44 | 45<br>(22 to 60) | 0.0104 |
| Plasma nitrite concentration (nM) | 570 (85) | 109 (12) | 461<br>(269 to 653) | 308 | 324<br>(125 to 441) | 0.0214 |
| Systolic blood pressure (mmHg) | -7 (2) | 1 (1) | -7<br>(-11 to -3) | 6 | 7<br>(3 to 9) | 0.0196 |
| Diastolic blood pressure (mmHg) | 5 (3) | 1 (1) | -6<br>(-9 to -2) | 2 | 3<br>(-4 to 6) | 0.5107 |

Data for the analysis involved 60 experimental conditions in $n = 15$ males. Systolic and diastolic blood pressure values were obtained immediately prior to, and 2.5 hours following, nitrate or placebo supplementation and data reflect the change in pre-to-post intervention values. Blood samples were obtained 2.5 hours post supplementation only, therefore data for plasma nitrate and nitrite concentrations reflect post-supplementation values only.
<sup>1</sup>Estimate 1: A naive estimate of the individual differences SD using the simple equation of $SD_{IR} = \sqrt{SD_S^2 - SD_P^2}$ where $SD_{IR}$ is the SD of the true individual response, and $SD_S$ and $SD_P$ are the SDs of the primary outcome measures in the supplementation and placebo conditions, respectively [23]
<sup>2</sup>Estimate 2: Individual differences SD estimated using a random effects within-subjects statistical model [25]. The SD was calculated from the participant-by-condition interaction term modelled as a random effect and the *P* value is for this interaction term.
SE, standard error; SD, standard deviation; CI, confidence interval; *int*, participant-by-condition interaction.

Linear mixed models revealed a main effect of condition (p<0.001), with a larger mean plasma nitrate concentration of 225 uM (95%CI: 199 to 250 uM) in the nitrate supplementation *versus* placebo condition. When averaged over the two replicates, the placebo-controlled post-supplementation mean plasma nitrate concentration ranged from 163 to 300 uM between the participants (Figure 2A). The meta-analytic approach-estimated between-participant replicate-averaged treatment effect nitrate response heterogeneity (τ) was ± 43 uM (95%CI: 30 to 69 uM).

**Figure 2.**
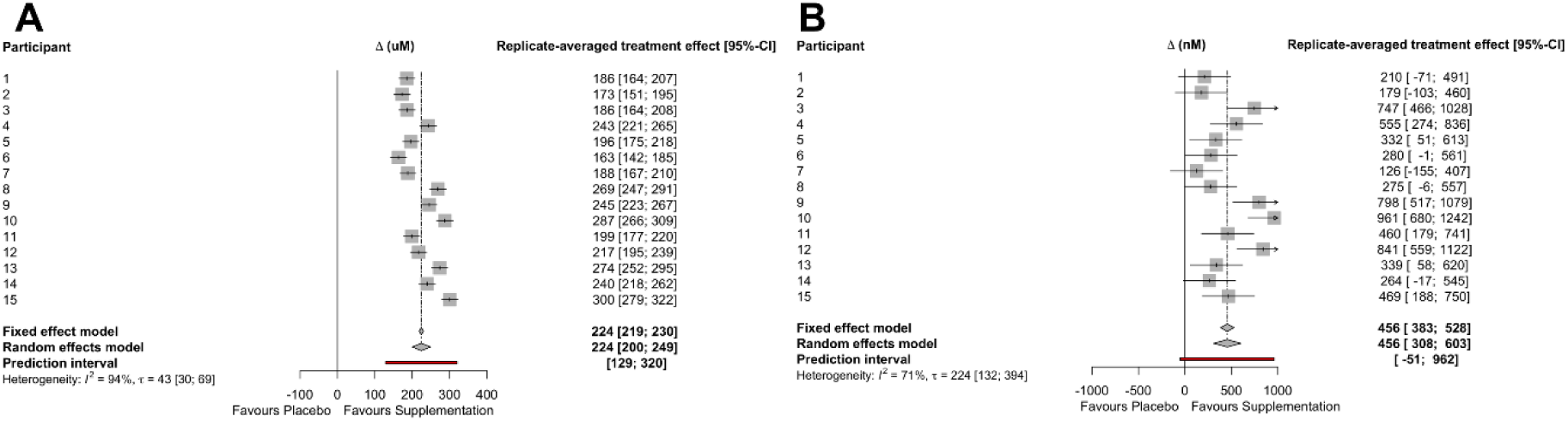
Forest plots for the plasma nitrate (A) and nitrite (B) summary effects and between-participant (t) replicated-averaged treatment effect heterogeneity. τ, denotes tau-statistic; CI, denotes confidence interval.

### Plasma nitrite concentration

There was a moderate positive correlation (r=0.55, 90%CI: 0.14 to 0.80) between the two sets of placebo-adjusted responses to dietary nitrate supplementation for plasma nitrite concentration (Figure 1B). The within trial SD for plasma nitrite concentration was substantially greater for the nitrate supplementation versus placebo conditions (Table 1), with the model-based treatment-by-condition interaction response variability of ±324 (95%CI: 125 to 441 nM). Linear mixed models revealed a significant main effect of condition (p<0.001), with the mean plasma nitrite concentration 461 nM higher (95%CI, 269 to 653 nM) in the nitrate supplementation versus placebo condition. When averaged over the two replicates, the placebo-controlled post-supplementation mean plasma nitrite concentration ranged from 126 to 961 nM between the participants (Figure 2B). The meta-analytic approach-estimated between-participant replicate-averaged treatment effect nitrite response heterogeneity (τ) was ± 224 nM (95%CI: 132 to 394 nM).

### Systolic BP

There was a strong positive correlation (r=0.80, 90%CI: 0.55 to 0.92) between the two sets of placebo-adjusted responses to dietary nitrate supplementation for systolic BP (Figure 1C). The within trial SD for systolic BP was substantially greater for the nitrate supplementation *versus* placebo conditions, and the model-based treatment-by-condition interaction response variability was ±7 mmHg (95%CI: 3 to 9 mmHg) (Table 1). Linear mixed models revealed a main effect of condition (p=0.001), with a mean reduction in systolic BP that was 7 mmHg (95%CI: 3 to 11 mmHg) greater in the nitrate supplementation *versus* the placebo condition. When averaged over the two replicates, the placebo-controlled nitrate supplementation response ranged from a 9 mmHg increase to a 24-mmHg reduction between the participants (Figure 3A). The meta-analytic approach revealed the upper confidence limit for the between-participant replicate-averaged treatment effect heterogeneity of τ = ± 7 mmHg (95%CI: 5 to 12 mmHg) surpassed the clinically relevant target reduction of 2 mmHg for 8 participants (Figure 3A).

**Figure 3.**
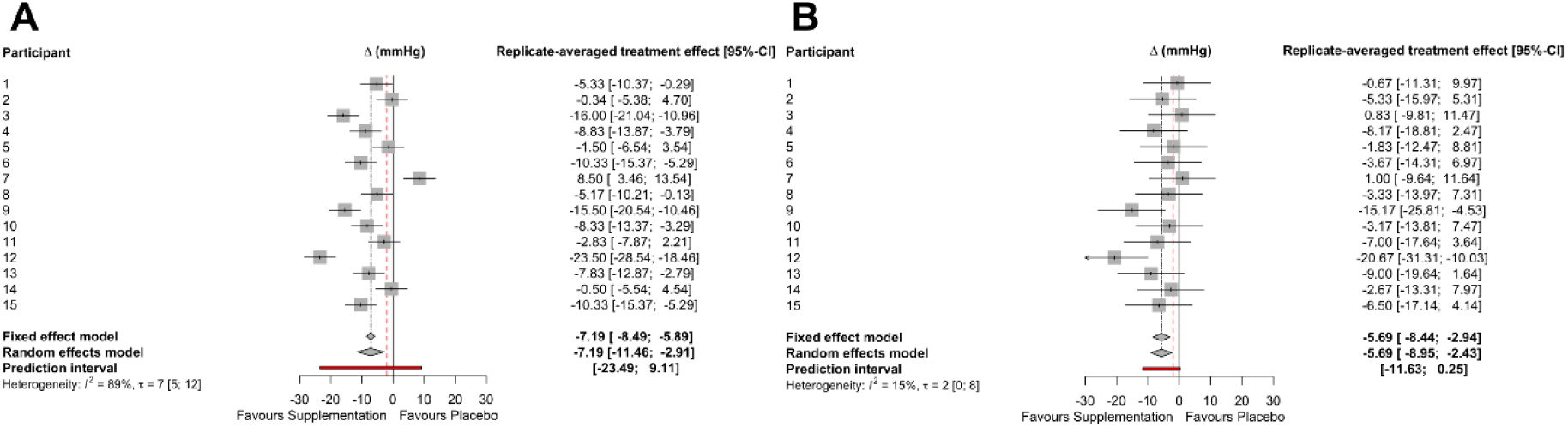
Forest plots for systolic BP (A) and diastolic BP (B) summary effects and between-participant (t) replicated-averaged treatment effect heterogeneity. The red dashed line indicates the target difference reduction of 2 mmHg. τ, denotes tau-statistic; CI, denotes confidence interval.

### Diastolic BP

Correlations between the two sets of placebo-adjusted responses to dietary nitrate supplementation for diastolic BP were small and non-significant (r=0.06, 90% CI: –0.39 to 0.49, Figure 1D). The within trial SD for diastolic BP was similar for the dietary nitrate versus placebo conditions, with the model-based treatment-by-condition interaction response variability of ±3 mmHg (95%CI: –4 to 6 mmHg; Table 1). Linear mixed models revealed a significant main effect of condition (p=0.003), with a mean reduction in diastolic BP of 6 mmHg lower (95%CI: 2 to 9 mmHg lower) in the nitrate supplementation *versus* placebo condition. When averaged over the two replicates, the placebo-controlled nitrate supplementation response ranged from a 1 mmHg increase to a 21-mmHg reduction between the participants (Figure 3B). The meta-analytic approach revealed the upper confidence limit for the between-participant replicate-averaged treatment effect heterogeneity of τ = ± 2 mmHg (95%CI: 0 to 8 mmHg) surpassed the clinically relevant target reduction of 2 mmHg for 2 participants (Figure 3B).

### Correlations amongst outcome variables

There was a small, non-significant positive correlation (r=0.37, 90%CI: –0.09 to 0.70, p=0.09) between the placebo-adjusted plasma nitrate and nitrite concentrations following dietary nitrate supplementation. In addition, there were small, non-significant negative correlations between the placebo-adjusted plasma nitrate concentrations and the change in both systolic (r=-0.20, 90%CI: –0.59 to 2.63, p=0.23) and diastolic (r=-0.26, 90%CI: –0.63 to 0.21, p=0.174) BP following nitrate supplementation. There was a moderate, significant negative correlation between the placebo-adjusted plasma nitrite concentrations and the change in both systolic (r=-0.76, 90%CI: –0.90 to –0.47, p<0.001) and diastolic (r=-0.51, 90%CI: –0.77 to –0.10, p=0.03) BP following nitrate supplementation. The slope of the regression line between the placebo-adjusted plasma nitrite concentration and the change in systolic (y=-0.016+1.791) and diastolic (y=-0.007+3.319) BP was –0.016 (95%CI: –0.022 to – 0.010) and –0.007 (95%CI: –0.016 to 0.001), respectively. Supplementary Table 1 illustrates the expected BP changes for a given plasma nitrite concentration.

## DISCUSSION

This is the first study on the topic of dietary nitrate supplementation to follow a replicate crossover design and associated expert guidance for data analysis (22, 25, 34). We detected inter-individual differences in the effects of dietary nitrate supplementation on plasma nitrate and nitrite concentrations and systolic BP that were distinguishable from random within-subject variability. In addition, our data suggest that the effects of nitrate supplementation on plasma nitrate and nitrite concentrations, and systolic BP are consistent within individuals when measured on at least two occasions.

We observed a mean reduction in systolic BP following dietary nitrate supplementation of 7 mmHg. A novel finding of our study is that the degree of systolic BP reduction following nitrate supplementation, i.e., the treatment response was highly variable between individuals, and greater than the within-participant trial-to-trial random variation. Our study, therefore, supports the notion that there may be genuine ‘*responders*’ and ‘*non-reponders*’(or, more accurately, higher and lower responders) to dietary nitrate supplementation [17–20]. Application of a recently described meta-analytic approach for replicate crossover trial examination revealed that the upper confidence interval for the control-adjusted BP reduction following nitrate supplementation exceeded the MCID of 2 mmHg for approximately half (8/15) of our participants. This suggests that these individuals are likely to experience BP reductions with nitrate supplementation of a magnitude which could potentially contribute to mitigating CVD incidence and mortality [38, 53].

Nitrate supplementation also significantly reduced diastolic BP, with a mean reduction of 6 mmHg. However, the between-replicate correlation for diastolic BP was low. This suggests that the effects of nitrate on diastolic BP are not as clear (relative to the random within-subjects variability) compared with systolic BP. This may help explain why reductions in diastolic BP are less frequently observed in the extant literature [7]. The participant-by-condition interaction was also small, suggesting inconsistent inter-individual variability in the effects of nitrate supplementation on diastolic BP. It is currently unclear why there appears to be repeatable interindividual differences in the effects of nitrate supplementation on systolic, but not diastolic, BP. It is possible that there are measurement or experimental issues that could obscure true inter-individual differences in diastolic BP responses, but more research is needed to explicate this phenomenon.

The potential for evidence-based personalised recommendations around dietary nitrate intake rests on formal identification of individual participants who will benefit most from consumption of this compound [54]. This could be achieved by identifying participant characteristics that are associated with the level of BP response to dietary nitrate. At the group level, previous data suggests that nitrate supplementation may be more effective at lowering BP in males versus females (although more studies in females are needed to confirm this hypothesis) [15, 55] and in younger versus older adults [56, 57]. All participants in our study were healthy young males which emphasises the importances of factors other than age, sex and health in determining inter-individual responses to nitrate supplementation.

Following consumption, dietary nitrate is absorbed in the upper gastrointestinal tract, increasing plasma nitrate concentrations [10]. Whilst most of the ingested nitrate is excreted in urine, approximately 25% is returned to the oral cavity via the salivary glands [58] where it is reduced to nitrite by oral bacteria [59]. This nitrite is swallowed and partly converted to NO and other nitrogen oxides in the stomach. Some nitrite also reaches systemic circulation, where it can be reduced to NO in various tissue [10]. Investigation of factors that may explain inter-individual variations in response to nitrate supplementation could focus on key steps within the gastrointestinal tract (including actions of oral bacteria involved in the regulation of NO bioavailability [21, 60, 61] and stomach pH which influences non-enzymatic conversation of nitrate to NO and other reactive nitrogen oxides [62]). Interestingly, recent data from Willmott et al. [63] suggests that individuals with a greater oral nitrate reducing capacity achieve a larger reduction in diastolic BP after nitrate supplementation. In addition, research should further explore the impact of genetic variants (or other factors) that alter the biological activity of the proteins e.g., nitric oxide synthases (NOS) involved in production of NO from both nitrate and other dietary sources (e.g., L-arginine/L-citrulline). Hobbs et al. [17] found that nitrate was more effective at lowering diastolic BP in T carriers (compared with GG carriers) of the Glu298Asp polymorphism in the gene (*NOS3*) encoding eNOS [17]. To date, there has been no systematic investigation of these and other factors that may be causally responsible for inter-individual differences in the response to nitrate supplementation using appropriate designs such as replicate crossover studies. This is a priority for future research with potential to be an important exemplar for precision nutrition.

Strengths of this study include the adoption of a replicate crossover study design and the use of appropriate statistical approaches for quantifying between-participant outcome response variability. This extends previous studies that have explored the impact of nitrate on biological markers and/or BP in traditional crossover or parallel group designs [7] or with repeat administration of nitrate *but not* control/placebo arms [21], such that the participant-by-condition interaction could not be estimated. Undeniably, the repeated administration of treatment and placebo for derivation of the person by treatment interaction complicates the statistical analysis, but this is a necessary complication for appropriate study of this topic.

Unfortunately, previous researchers have arrived at erroneous claims about treatment response hetergeneity on the basis of simple, but compromised, responder counting and simply observing individual changes solely from the treatment group in a trial [23]. Potential limitations include only recruiting healthy young males as participants. While this may represent a limitation in generalisation of our findings to the general population, this design feature was a distinct advantage in revealing evidence of true inter-individual variations in physiological response to nitrate supplementation that cannot be explained by sex or age differences alone. Whilst lowering BP in this group may be less of a public health priority than individuals with hypertension, lowering BP in individuals who are not hypertensive down to a least ∼115/75 mmHg could reduce risk of vascular death [53]. A further limitation is the relatively low sample size (n=15), although both the crossover (within-subjects) and replicate aspects of our design increased statistical power (relative to a parallel arm study) for detection of mean treatment effects. Our design and sample size also enabled the detection of statistically significant individual differences in responses for all outcome variables apart from diastolic BP. We cannot rule out the possibility that response heterogeneity in diatolic BP would be detected with a larger sample size, more reliable measurements and/or a greater number of replicates in the design. Repeated administration of treatments is the key aspect for detecting, confidently, treatment response heterogeneity but laboratory-based and somewhat invasive replicate crossover studies like ours are difficult to recruit for. Additionally, we studied acute responses so it remains to be established whether there are similar inter-individual differences in response to nitrate supplementation over longer intervention periods. Longer-term interventions might also clarify whether the benefits of nitrate consumption on CVD risk proposed in observational studies [64] occurs with prolonged dietary nitrate supplementation, and whether benefits are restricted to certain population sub-groups/individuals.

This study revealed evidence of substantial inter-individual differences in the physiological responses (NO biomarkers and systolic BP) to dietary nitrate supplementation in healthy young males. This provides proof-of-concept as a basis for further investigations of the magnitude, durability and pervasiveness of these inter-individal responses across diverse populations and of the (biological) factors responsible for the observed intra-individual variation. These findings open up the exciting possibility of personalised recommendations for dietary nitrate intake for optimal management of BP and related health outcomes.

## Data Availability

Data described in the manuscript, code book, and analytic code will be made available upon request pending author approval

## ACKNOWLEDGEMENTS

We would like to thank our participants for giving up their time to take part in this study. In addition, we would like to thank Mr Jack Williams for assisting with the plasma nitrate and nitrite analyses.

## AUTHOR CONTRIBUTIONS

OMS, EH, JCM, MS, RK, LL, GA, EW, JM and AG designed the research. EW generated the random allocation sequence and ensured researchers involved in participant enrolment and data collection (OMS, EH and SA) were blinded to the experimental conditions. EH, SA, and OMS collected the data and MB and CE conducted the plasma nitrate/nitrite analyses. GA, LL and OMS conducted the statistical analysis. OMS, EH, SA, MS, EW, RK, JM, AG, MS, MB, CE, LL, GA and JCM critically interpreted the data. OMS, EH, SA, MS, EW, RK, JM, AG, MS, MB, CE, LL, GA and JCM wrote the paper. OMS had primary responsibility for the final content. All authors have read and approved the final manuscript.

