## Supplementary material for "Inter-individual differences in the blood pressure lowering effects of dietary nitrate: A randomised double-blind placebo-controlled replicate crossover trial"

**Supplementary Table 1.** Estimated marginal means for blood pressure changes by plasma nitrite concentration

| Plasma Nitrite (nM) | Systolic blood pressure (mmHg) | | | Diastolic blood pressure (mmHg) | | |
|---|---|---|---|---|---|---|
| | Δ | 95%CI | | Δ | 95%CI | |
| 200 | -1 | -3 | 2 | -1 | -4 | 2 |
| 300 | -2 | -5 | 0 | -2 | -4 | 1 |
| 400 | -4 | -6 | -2 | -2 | -5 | 0 |
| 500 | -6 | -8 | -3 | -3 | -6 | 0 |
| 600 | -7 | -10 | -4 | -4 | -7 | 0 |
| 700 | -9 | -12 | -5 | -5 | -9 | 0 |
| 800 | -10 | -14 | -7 | -5 | -10 | -1 |

**Δ, denotes expected blood pressure changes for a given plasma nitrite concentration; CI, denotes confidence interval.**

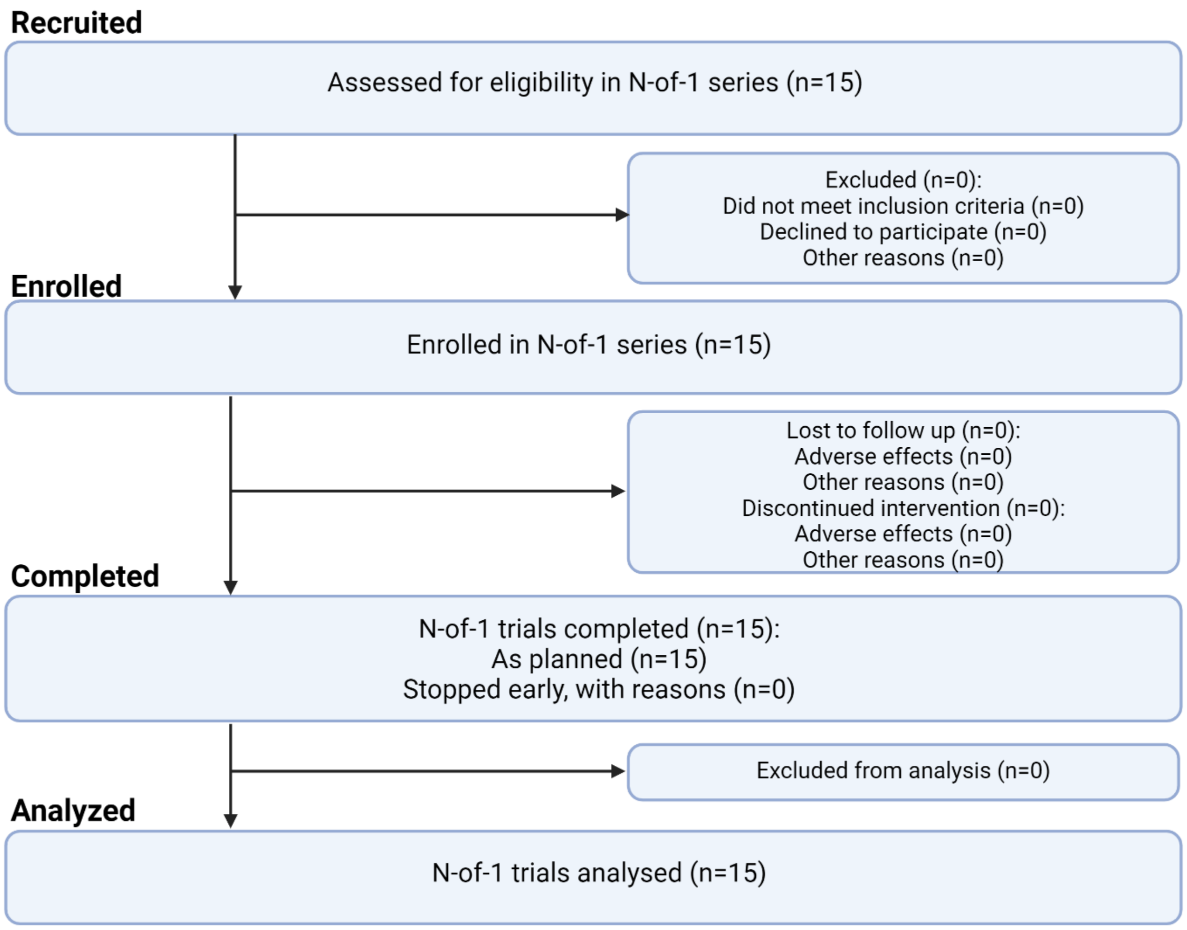


**Supplementary Figure 1.** Adapted CONSORT Flow chart according the CENT extension guidelines for n-of-1 trials

**Supplementary Text 1: Randomisation order**

**Source:** Generated using http://www.randomization.com

**Participant 1:** ABBA
**Participant 2:** BBAA
**Participant 3:** ABBA
**Participant 4:** ABAB
**Participant 5:** AABB
**Participant 6:** BAAB
**Participant 7:** BABA
**Participant 8:** BBAA
**Participant 9:** AABB
**Participant 10:** AABB
**Participant 11:** BAAB
**Participant 12:** BBAA
**Participant 13:** BABA
**Participant 14:** BABA
**Participant 15:** BAAB
